# Dementia Education and Training for the Multidisciplinary Student Healthcare Workforce: A Systematic Review

**DOI:** 10.1101/2025.01.15.25320583

**Authors:** Malvika Muralidhar, Saskia Delray, Claudia Cooper, Sedigheh Zabihi, Sube Banerjee, Clarissa Giebel, Karen Harrison-Dening, Yvonne Birks, Charlotte Kenten, Madeleine Walpert

## Abstract

**Objectives:** To systematically review existing policies and research evidence on the effectiveness of dementia education and training for health and social care students.

**Methods:** We searched electronic databases for primary research studies (published between 2015-2024), evaluating dementia training for health and social care students. We assessed risk of bias using the Mixed Methods Appraisal Tool, prioritising studies scoring 4+ (higher quality) that reported significant findings on primary outcomes from controlled intervention trials. We reported outcomes using Kirkpatrick’s framework. We consulted professional stakeholders in focus group regarding how findings might inform practice.

**Results:** 17/35 included studies were rated 4+ on the MMAT; only one met our criteria for priority evidence. An experiential programme for medical students, “Time for Dementia”, which combined skill-learning and reflective sessions with visits to people with dementia, was found to improve Kirkpatrick Level 2 (learning) outcomes, attitudes and knowledge over two years of participation; this was supported from findings from qualitative studies. Asynchronous, self-directed learning did not improve learning outcomes, relative to standard training. Though almost all training programmes sought to learn from lived experience, none consulted patients regarding the impact of the training on students’ skills and practice. Nine focus group attendees agreed that the evidence reflected their experiences that consistent support, combined with skills-based and reflective sessions, optimised student learning from initial patient-focused encounters.

**Conclusions:** Effective interventions increased confidence and enjoyment of dementia care encounters, and increased interest in careers in dementia specialties. Mandating evidence-based dementia skills programmes across specialties could ensure that students learn the skills and competencies required to be part of an effective future workforce and drive important improvements in care quality. Evidence based approaches to enhancing dementia education in training could include experiential learning modules in early years of medical school training and allied health and care professional training, using evidence-based approaches to teach communication skills and other essential dementia care skills within clinical placements, and providing dedicated supervision to support their implementation. Future research could usefully consider patient perspectives in determining the impact of educational programmes.

## Introduction

Over 900,000 people have dementia in the UK^1^. This is predicted to rise to 1.4 million by 2040 ^2^. Dementia is a leading cause of mortality and morbidity^3^. Nine in ten people with dementia have at least one comorbid long-term condition, therefore healthcare professionals across all specialties require the knowledge and skills to deliver high quality, compassionate dementia care^4,5^. Pre-qualification is the optimal time to deliver dementia education^6^, though there are concerns that current curricula do not adequately cover it, especially community dementia care^7^. Lack of preparedness and understanding of dementia can lead to high staff turnover and poor retention rates^8^.

Current training programmes vary in content and duration, with innovative training often limited to brief placements taken up by the keenest students^9^. The Dementia Core Skills Education and Training Framework (DCSETF) specifies learning outcomes and standards for curricula and commissioning of education programmes^10^. While several programmes have been developed which aim to enhance dementia care competencies, few are evidence-based^11^. This paper is part of a set of evidence reviews regarding how dementia skills and knowledge are best supported in the health and social care workforce, with evidence for knowledge and skill development in the current workforce published separately^12^. We aimed to systematically review evidence for dementia training programmes for health and social care workers before qualification. Our research questions were:

1. What is the current evidence on how dementia skills and knowledge are best taught to health and social care students (pre-qualification) to prepare them for practice?
2. How feasible and useful do stakeholders consider the existing evidence to implement?

## Methods

### 2.1 Systematic review

The study was conducted in line with the Preferred Reporting Items for Systematic Reviews and Meta-Analyses (PRISMA; Figure 1) recommendations^13^ and registered in PROSPERO (CDR42024509026).

**Figure 1:**
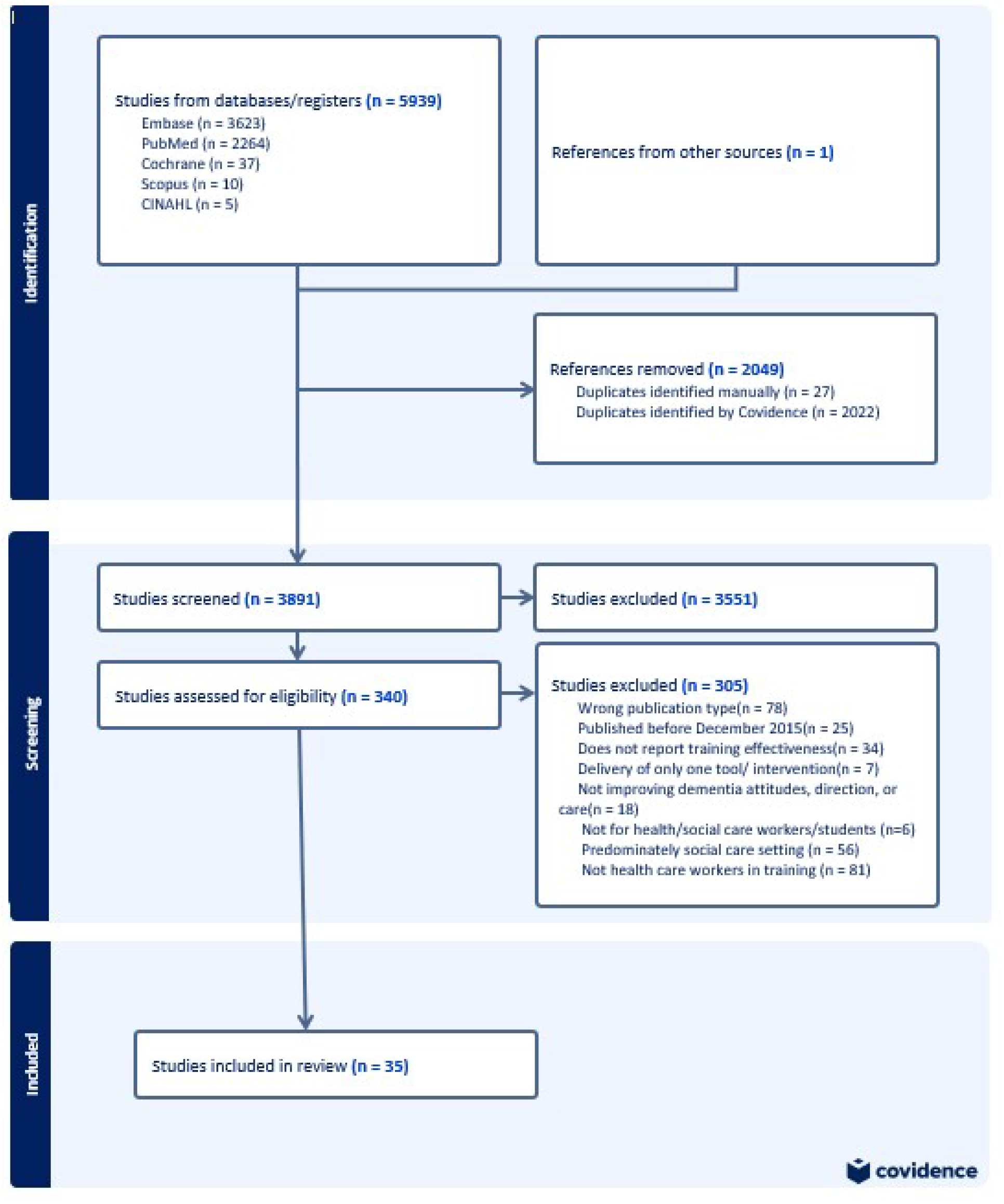
PRISMA (Preferred Reporting Items for Systematic Reviews and Meta-Analyses) diagram of included and excluded studies.

#### 2.1.1 Search strategy

We systematically searched electronic databases including PubMed, Embase, Scopus, CINAHL and the Cochrane Library from 01 December 2015 (updating previous synthesis^14^) to 20 February 2024. Terms related to ‘education’/ ‘training’, ‘staff’, and ‘dementia’ were combined with Boolean operators ‘AND’ and ‘OR’ linking search terms within concepts (Table S1). We asked experts in the field to identify unpublished studies and searched references of included papers for additional studies.

#### 2.1.2 Eligibility criteria and study selection

Following title and abstract screening, two reviewers (SD, CC) independently screened full-text papers against criteria to identify eligible studies. Discrepancies were resolved through discussion. We included primary (quantitative, qualitative and mixed method) research studies, reporting the effectiveness of any training or educational intervention focused on developing dementia-specific knowledge, values and skills, for health care or social work students (prior to award of a license to practice). We excluded individual case studies, dissertations and meeting abstracts.

#### 2.1.3 Data Extraction and Quality Appraisal

We developed a standardised form to extract data from included studies, describing characteristics of students recruited, interventions and control conditions (where relevant); outcomes and response rates (Table 1). We used Kirkpatrick’s model to classify whether evaluated interventions demonstrated effectiveness in terms of: learner’s reaction to and satisfaction with (Level 1); knowledge, skills, confidence, and attitude change indicating learning has occurred (Level 2); change in student behaviour or practices (Level 3); and attainment of targeted outcomes (client wellbeing: Level 4) ^15^.

**Table 1:**
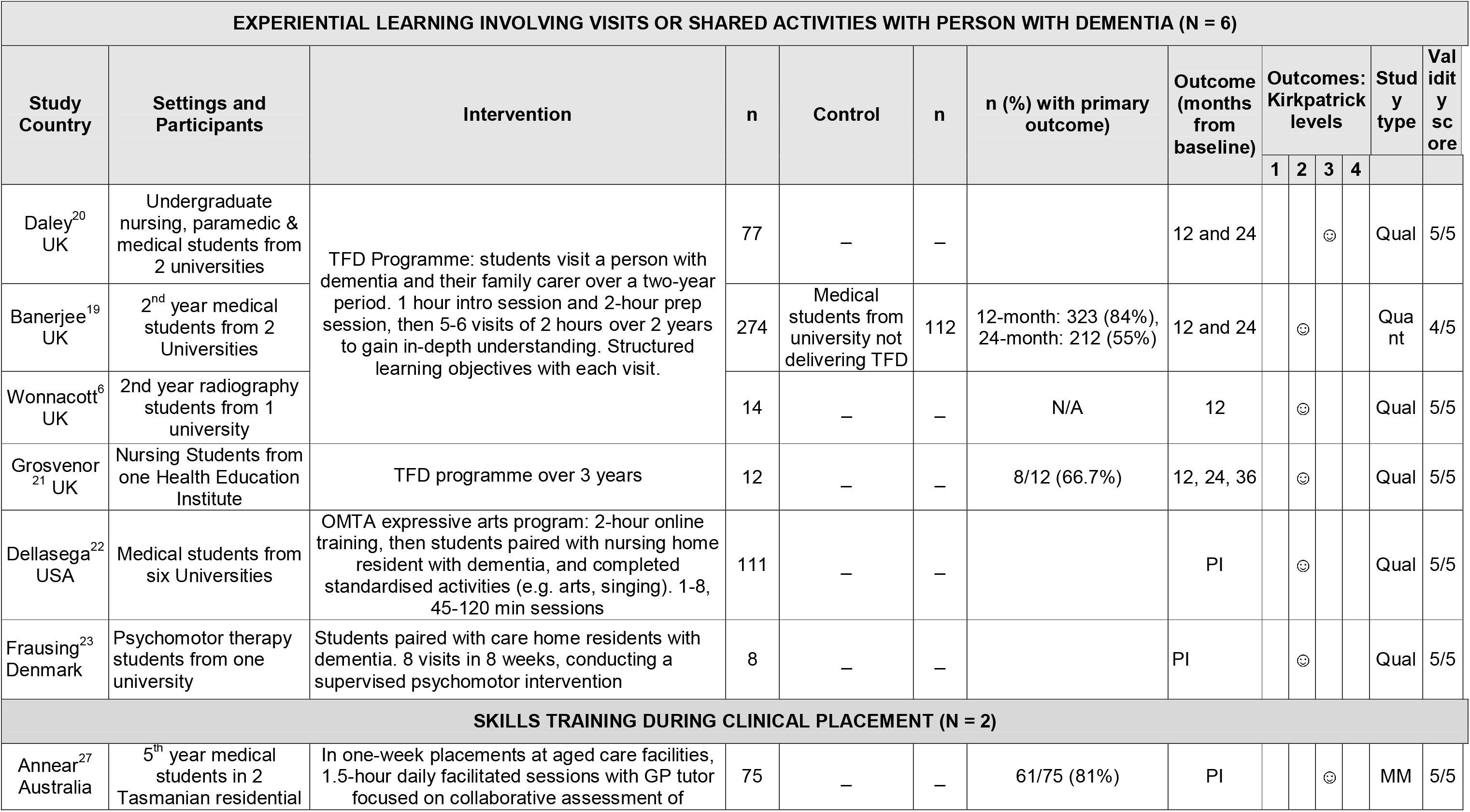
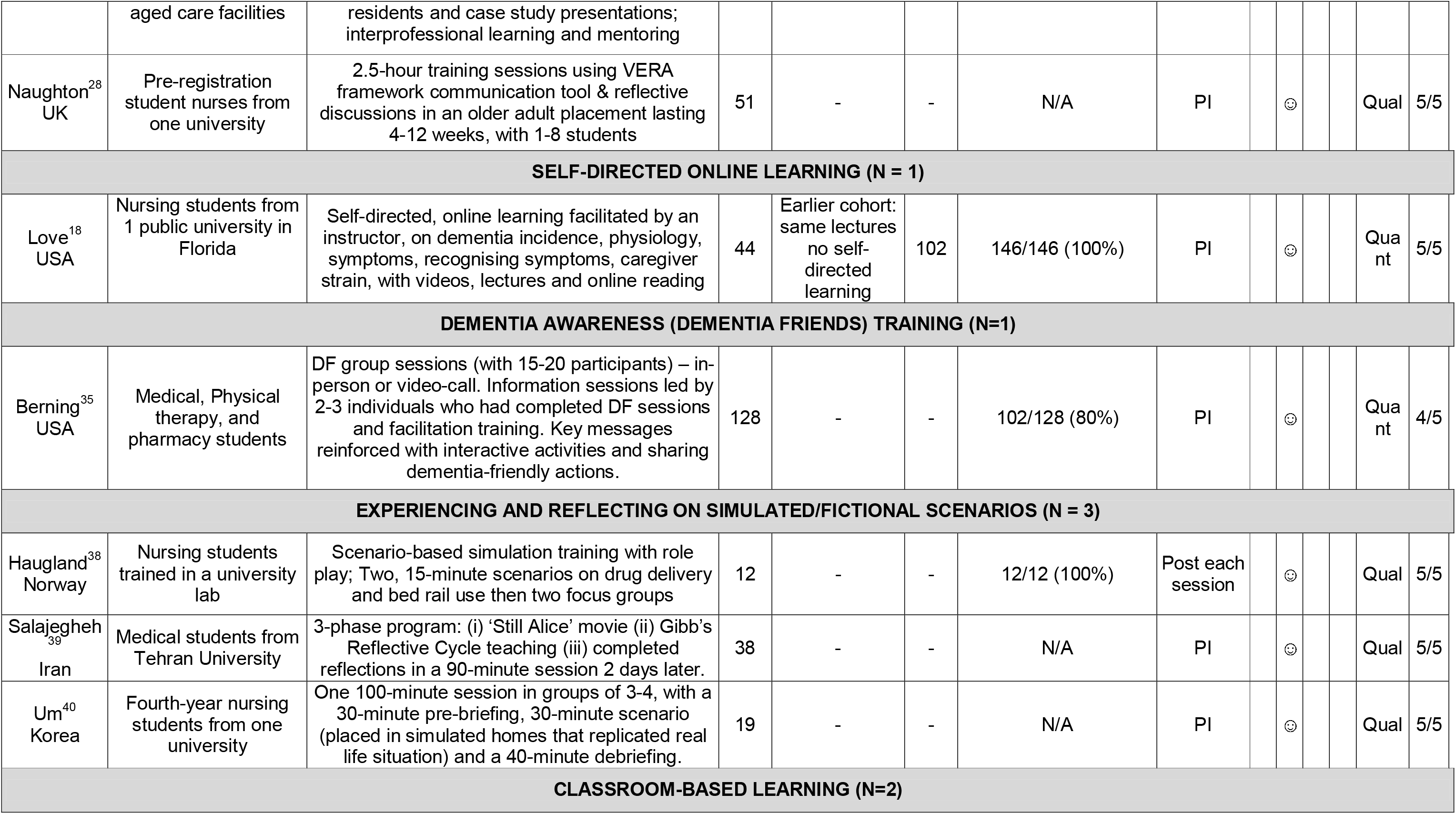
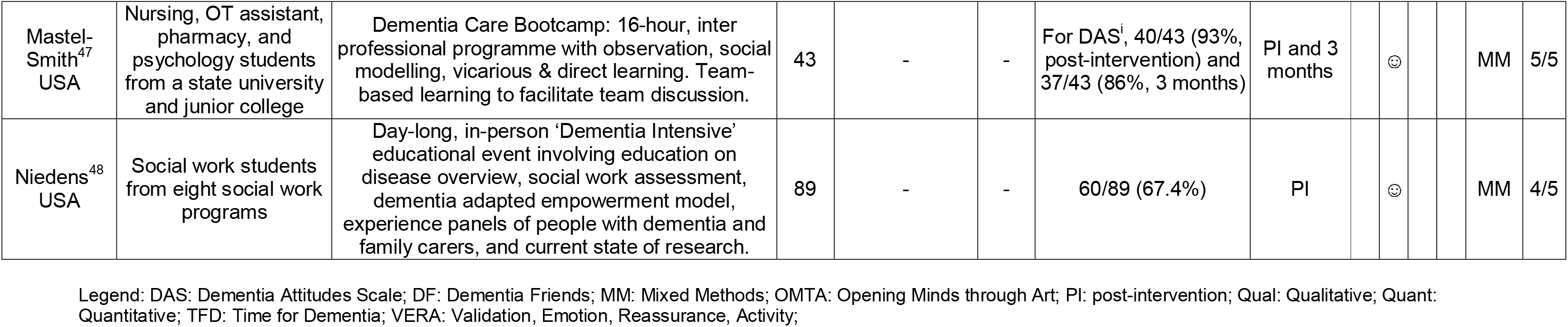
Characteristics of studies rated 4+ on the MMAT appraisal tool.

SD, SZ, MM and MW independently assessed the quality of included studies using the Mixed Methods Appraisal Tool^16^. Two reviewers independently rated the studies and resolved disagreements in discussion with a third reviewer (Table S2). Studies were scored out of five; a rating of four or above was considered high quality^12^.

#### 2.1.4 Data Analysis

MM and CC independently categorised studies by intervention types, then discussed with other authors to determine final categorisations. Within each category, we narratively synthesised findings, mapping reported outcomes to Kirkpatrick’s framework^15^. We focused on evidence from higher quality studies (predefined as score 4+), and within these ascribed the label ‘priority evidence’ to studies that reported a significant finding on a quantitative main outcome in a between-group comparison (intervention versus control condition).

### 2.2 Stakeholder consultation

To answer RQ2, we held an online workshop in November 2024, inviting care professionals from diverse backgrounds, seniorities and roles identified via DeNPRU-QM national networks. MM presented findings from evidence-supported interventions to facilitate discussions on which type of intervention would be most appropriate to implement in practice and for learners’ needs. SZ chaired, and MM noted discussions verbatim. CK, SD, and CC were also present. SZ and MM met after the workshop to discuss and synthesize findings from discussions and identify key themes. These were circulated to attendees, who were invited to comment further.

## Results

### 3.1 Systematic review

We identified 5,939 studies in our electronic search, of which 35 met inclusion criteria (Figure 1 shows search results).

### 3.1.1 Study characteristics

Included studies used non-randomised quantitative (n=13), qualitative (n=10), and mixed methods (n=12) designs. They evaluated educational and training interventions for students of nursing (n=13), from more than one discipline (n=9), medicine (n=7), with single studies evaluating training for psychomotor therapy, paramedic, pharmacy, social work, physiotherapy, and radiography students.

17/35 (48.5%) of included studies were rated as high quality (4+ on the MMAT). Based on the content of interventions, we categorise studies as:

1. Experiential learning (visits with people affected by dementia, beyond usual clinical placements)
2. Additional skills training and reflective sessions during usual clinical placements
3. Self-directed online learning
4. Dementia awareness (Dementia Friends) training
5. Learning from simulated/fictional scenarios
6. Classroom-based learning activities.

We summarise studies scoring 4+ on the MMAT (higher quality) in Table 1 and prioritise them in our narrative summary below; summarising studies scoring <4 on the MMAT in Table S3. Only two higher quality studies included a control group^17,18^, of which the first reported a statistically significant finding on a primary outcome and so met our criteria for priority evidence.

#### 3.1.2 Experiential learning (n=9)

Five of these studies evaluated the UK Time for Dementia (TFD) programme. In TFD, after an initial meeting covering programme aims, student expectations, safeguarding and communication skills, pairs of students visit a person with dementia and their family for two hours several times a year, usually over two years. Course handbooks guide student visit activities, including: discussing how dementia affects the person and family, and how they experience health and social services; using reminiscence and storytelling as enjoyable and empowering activities; exploring client care needs and preferences. Students complete a reflective assignment and attend a final conference with all students, people with dementia and carers participating in the programme^19^.

##### High quality evidence (n=6)

We found high quality evidence to support TFD. The only study that met our criteria for prioritisation was a non-randomised, parallel group comparison of second-year students from two medical schools, one where the TFD programme was compulsory and one (control group) where it was not taught^19^. Adjusting for student age, gender, and previous dementia experience, relative to the control group over two years, students who received TFD reported improved attitudes to dementia (primary outcome) on the Approaches to Dementia Questionnaire (ADQ) (Coefficient: 2.19, 95% confidence interval [95% CI]: 0.75– 3.64, p =, Dementia Knowledge Questionnaire (DKQ) scores (1.63, 1.04–2.23, p < 0.001) and Dementia Attitudes Scale (DAS) (6.55, 3.91–9.19, p < 0.001), with no change on empathy measures.

Three single-group qualitative studies explored experiences of TFD among participating medical, nursing and paramedic^20^; nursing^21^ and radiography^6^ students. Students indicated they learned and used communication skills (e.g. speaking slowly, checking for understanding), and becoming mindful of taking time to hear family members^20^. A fourth qualitative study explored the impact of Open Minds through Art (OMA), a programme for USA medical students. Similar to TFD, OMA included an initial (two-hour online) induction, then paired students with a person with dementia (in this programme, in residential care) to engage in standardised activities during visits (such as visual artwork and singing familiar songs) and reflective essay coursework^22^. Students completed between 1-8 sessions of up to two hours. A fifth qualitative, Danish study evaluated an extracurricular programme for undergraduate relaxation and psychomotor therapy students. They visited a person with dementia in a care centre weekly for eight weeks, conducting an intervention similar to gentle massage ^23^. Induction meetings introduced dementia care and the specific intervention. The intervention was supervised by a psychomotor therapist, but did not appear to include any formal reflective activity, aside from a research focus group. All five studies explored themes of improved communication skills, increased confidence to overcome challenges and re-evaluating negative dementia attitudes and stereotypes.

##### Lower quality evidence (n=3)

A further TFD study echoed the findings of higher quality studies^24^. Pharmacy students rated their comfort level and perceived communication abilities higher on an unvalidated survey, after an experiential intervention in which they spent time supporting long-term care facility residents with dementia to derive benefit from fidget blankets^25^. A German study reported that a 30-hour, inter-professional training course (for medical students and student nurses) comprising introductory tutorials and care home visits in which skills were applied^26^ was acceptable and feasible.

##### Summary of evidence

- There is Kirkpatrick Level 2 evidence from a high quality, controlled study that Time for Dementia, an experiential programme for medical students with skill-building and reflective sessions, improved dementia knowledge and attitudes during the two years of the programme. Qualitative, higher quality studies consistently supported the value of similar programmes for medical students and allied health professionals in training.

#### 3.1.3 Skills training during clinical placement (n=5)

##### High quality evidence (n=2)

Two single-group, higher quality studies evaluated programmes to enhance clinical students learning during clinical placements. An Australian mixed methods study evaluated an innovative, mandatory clinical placement for fifth year medical students in residential aged care facilities^27^. Students assessed volunteer residents, collaboratively with facility nursing and care staff, and presented cases within daily, 1.5-hour supervision groups with a GP tutor, where they were encouraged to critically analyse clinical issues and make treatment and care recommendations. 61/75 (85%) of placement students completed outcome measures, students’ Dementia Knowledge Assessment Tool (DKAT 2) scores (*z* = -2.63, *p* = 0.009, *r* = 0.37) and their enjoyment working with older people (*z* = -3.08, *p* = 0.002, *r* = 0.27) increased post-placement. In focus groups, students described being empowered by greater appreciation of residents’ complex health issues, to address problems they encountered. They valued opportunities to provide new diagnoses or treatment plans.

A UK, qualitative study tested training based on the Validation, Emotion, Reassurance, Activity (VERA) framework developed to teach effective communication skills to use with people with dementia ^28^. VERA guides the exploration of unmet needs and supports staff in developing therapeutic relationships. They evaluated a 2.5-hour face-to-face training session which 51/64 (80%) of eligible students attended at the start of their older adult placement. Learning was revisited during the placement in reflective discussions led by clinical lecturers. Students described valuing opportunities to practice using the framework in role-play, and feeling they had increased their knowledge and understanding of dementia communication approaches.

##### Lower quality evidence (n=3)

Two lower quality studies also evaluated the VERA framework, one using the same cohort as the higher quality study above. Relative to a usual training comparison group, intervention group students were more likely to identify person-centred responses from bespoke case vignettes (the primary outcome)^29^. An Irish qualitative study allocated second-year nursing students to either receive VERA framework-based communication training or standard communication skills^30^. Themes identified in focus groups for both studies were feeling supported to recognise patient’s emotional needs, and the framework building confidence in providing care. Finally, a study explored the impact of a single workshop for student nurses during a geriatric hospital placement in which self-reflection, small group activities, case studies, and in-house developed simulation videos were used to teach dementia care and communication skills, with the aim of enhancing placement learning. Attendees felt a higher sense of efficacy and competency after the workshop ^31^.

##### Summary of evidence

- Two single-group, higher quality studies found that additional skills-building and reflective sessions during clinical placements increased knowledge acquisition in medical and nursing students (Level 2 evidence). There was preliminary Level 3 evidence that such sessions (involving an interdisciplinary element in a care home setting) helped medical students participate more actively in patient management.

#### 3.1.4 Self-directed online learning (n=4)

##### High quality evidence (n=1)

In the second of only two quantitative studies we identified to include a control condition, a US study compared traditional lecture-based teaching received by an earlier cohort of 102 undergraduate nursing students, with the same teaching plus self-directed online learning in the current cohort of 44 students, comprising at least four of 25 one hour e-learning modules on the USA National Institute of Aging website, on topics related to dementia diagnosis and management, caregiver support and managing challenging behaviours^18^. Adherence was not measured. Scores on the primary outcome of the Basic Knowledge of Alzheimer’s Disease (BKAD) increased in both groups, but post-intervention between group comparisons did not favour the intervention.

##### Lower quality evidence (n=3)

A Northern Irish, single-group study evaluated access to a co-designed, digital game^32^ in which players respond to multiple-choice questions testing dementia knowledge, attitudes, and behaviours. The game lasted around 90 seconds, and participants could play multiple times over four weeks. Adherence was not reported. Scores on all ADKS subscales increased post-intervention. Two US, single-group studies evaluated the impact of online modules, both incorporating videos and case vignettes, on nursing students’ dementia knowledge. Scores on dementia knowledge increased in the first study^33^ as did Sense of Competence in Dementia Care Staff (SCIDS) scores, but knowledge scores did not in the second study^34^.

##### Summary of evidence

- The only (high quality) study to include a comparison group did not find a difference between standard lecture format and lectures with additional online learning, suggesting that self-directed learning may not significantly enhance knowledge acquisition, relative to standard training.

#### 3.1.5 Dementia awareness (Dementia Friends) training (n=3)

Three studies evaluated one hour, dementia awareness workshops, with learning objectives that mapped to Tier 1 of the DCSETF^10^.

##### High quality evidence (n=1)

A single-group, US study evaluated a Dementia Friends workshop (one group in person, others online) for medical, physical therapy and pharmacy students. The sessions were scripted and used a workbook with interactive activities and communication tips, ending with encouragement to commit to one dementia friendly action ^35^. DAS dementia knowledge (p < 0.001), and comfort (p < 0.001) scores increased post-session.

##### Lower quality evidence (n=2)

A Malaysian study explored a one-hour dementia awareness initiative comprising demonstrations, interactive team activities, videos, discussions, and a case study^36^. They found that pharmacy and medical student attitudes towards people with dementia significantly improved post-workshop. Second, a UK study developed and qualitatively evaluated a learning package for students to gain a Tier 1 Dementia Awareness qualification^37^. First-year healthcare students from diverse disciplines attended a one-hour Dementia Friends session (face-to-face or online); completed a reflective workbook and answered multiple-choice questions. Post-intervention surveys found that 95% of participating students found this method of obtaining the qualification helpful.

##### Summary of evidence

- A Dementia Friends awareness course improved attitudes and knowledge immediately post-intervention (Kirkpatrick Level 2), in a single-group, higher quality study.

#### 3.1.6 Experiencing and reflecting on simulation and fictional scenarios (n=9)

##### High quality evidence (n=3)

Three qualitative studies evaluated training interventions involving experiences of simulation and fictional scenarios. In Norway, a study explored the effect of simulation training on first-year nursing students. 12/12 (100%) students invited completed the course, with scenarios involving drug delivery and using bed rails when caring for people with dementia^38^. An Iranian study tested an interactive programme for medical students based on watching the ‘Still Alice’ movie, about a woman with Alzheimer’s disease^39^. Finally, Um (2023) explored Korean nursing students’ experiences of simulated community visits. Student groups of 3-4 took part in 100-minute sessions with a pre-briefing, scenario, and a debrief. Similar themes emerged across these studies in post-intervention qualitative interviews and focus groups: increased empathy, a more holistic view of the circumstances of people with dementia and self-reflection on values and attitudes.

##### Lower quality evidence (n=6)

Three lower quality studies used Virtual Reality to simulate clinical scenarios, with the aim of increasing empathy. A Chinese study investigated the effect of watching the movie, ‘Still Alice’, followed by an eight-minute Virtual Dementia Tour (VDT) on nursing students^41^. Empathy levels increased post-test. Second, a US study investigated VR simulation as a learning tool for second year, pre-clinical medical students^42^. The 30-minute simulation was through the perspective of a middle-aged Latina woman diagnosed with AD, who the learner embodies, experiencing her struggles engaging with the public and family life and communicating to her family that she is confused. The simulation includes altering visual and auditory processing. Compared to pre-survey responses, there was an increase in students’ understanding of the effects of dementia (p<0.001) and the needs of people with dementia and their families. Finally, a US study explored nurse practitioner students’ experiences of a virtual reality intervention^43^. They found a significant improvement in scores of perception and reactions to dementia challenges. The key qualitative theme was developing empathic understanding.

Three other low quality studies simulated clinical scenarios in person. A UK study conducted twelve simulation days over nine months, based on communicating with a person with dementia in various scenarios^44^. Medical students undertook the simulation days, comprising ten-minute scenarios and 30-minute debriefs, facilitated by doctors, nurses, and occupational therapists specialised in dementia. Learners felt more confident about managing situations that initially felt uncomfortable, simulations challenged assumptions and increased students’ awareness of the language they used while talking about and interacting with people with dementia. Another UK study evaluated the effectiveness of the ‘Sliding Doors’ educational intervention^45^. Third-year social work and nursing students attended a one-day workshop that combined interactive drama-based learning, theatre, and participatory learning. Only social work students showed significant attitudinal shifts on an unvalidated measure. Qualitative findings indicated tensions and different perspectives between nursing and social work students. Finally, a two-hour experiential learning activity, comprising a simulated dementia experience, education, observation and debriefing did not improve physical therapy students’ attitudes towards people with dementia ^46^.

##### Summary of evidence

- There was high quality evidence from three, single-group qualitative studies that simulated scenarios of challenging situations and day-to-day life of a person with dementia (with pre-briefing and debriefing activities) changed Kirkpatrick Level 2 outcomes including attitudes, empathy and confidence of medical and nursing students in providing person-centred care, immediately post-intervention.

#### 3.1.7 Classroom-based learning (n=5)

##### High quality evidence (n=2)

A US study evaluated a dementia care bootcamp, a 16-hour team-based, interprofessional programme for healthcare students ^47^. Learners were placed in interprofessional teams consisting of nursing, pharmacy, and psychology students, and occupational therapy assistants. Students were sent pre-reading content. Bootcamp content was delivered by a dementia expert facilitator and included videos, case vignettes, lived experience, team-based role play, and a dementia virtual reality simulation, in which students completed five activities while garbed so that their vision, hearing, and sensations were impaired. This was followed by debriefing. Adherence was not measured. DAS (p < 0.001), DKAT 2 (p < 0.001), Confidence in Dementia (CODE) scale (p < 0.001) and perspective taking subscale of the Interpersonal Reactivity Index (IRI) (p < 0.05) improved post-test. Three months after baseline, students scored significantly lower on attitudes but significantly higher on knowledge, confidence, and empathy scales compared to pre-test (all p < 0.001). Themes from qualitative findings included valuing interprofessional learning. A second USA, single-group study evaluated a day-long “dementia intensive” for social work students, social workers and other professionals^48^. Dementia Knowledge Assessment Scale (DKAS) scores (mean difference = 9.9) and negative attitudes towards dementia (10% lower, p<0.05) improved post-training.

##### Lower quality evidence (n=3)

Three lower quality studies evaluated classroom-based learning programmes lasting one to three days. A German three-day workshop similarly focused on patient-centred care and interprofessional collaboration, using case-based learning^49^. Physiotherapy, nutrition therapy and counselling, and speech-language pathology students who attended the workshop significantly improved on the University of the West of England Interprofessional Questionnaire (UWE-IP-D) and provided positive feedback that the workshop was helpful in improving multidisciplinary collaboration.

Another US study evaluated a Student Ambassador program, comprising six educational and three outreach events, including monthly dementia-focused didactic meetings^50^. Medical students, undergraduate students and graduate students interested in medicine completed the program and showed significant improvements in knowledge, empathy and attitudes. Finally, a UK study evaluated a six-hour education programme on dementia for paramedic students, comprising didactic teaching by a dementia educator, covering subjects from the DCSETF and quizzes, case studies, videos, and pre-requisite learning^51^. Students showed a significant increase in knowledge and confidence in caring for someone with dementia after the sessions.

##### Summary of evidence

- There was Level 2 evidence from two single-group, higher quality US studies that a two-day dementia interprofessional “bootcamp” and a one day “intensive” incorporating a range of learning methods (interactive, simulation, learning from lived experience) improved knowledge, confidence, and empathy post-intervention and in one study for up to three months.

### 3.2 Stakeholders consultation

We consulted eight healthcare professionals from GP, psychiatry (n=2), nursing (n=3), clinical psychology, occupational therapy (OT) backgrounds, and one medical student. The group reflected on the value of interventions that might enhance learning on clinical placements and patient-focused encounters, with a nurse educator remarking that the VERA framework appeared easy to apply. Self-directed learning was considered of limited value in teaching communication and interpersonal skills, with a GP stating that practical teaching (e.g., real case discussions) is more useful. A medical student commented that learning in placements depended on the skills of the clinicians they were placed with and so varied and that some structure could increase consistency of placement experiences.

## Discussion

We found a paucity of evidence for dementia training for health and social care students. Considering the importance of equipping the 1.3 million-strong NHS workforce^52^ and social care workers with dementia knowledge and skills, the lack of high quality evidence on how best to deliver it effectively is striking. The only study meeting criteria for priority evidence found that an experiential programme for medical students (Time for Dementia) combining skill-learning and reflective sessions with patient visits, improved Kirkpatrick Level 2 (learning) outcomes, attitudes and knowledge over the two year programme. In single-group studies, we found consistent evidence that learning activities for medical and nursing students embedded in clinical encounters (usual placements, experiential visits or simulated encounters), including sessions to teach critical skills and for reflection, improved Level 2 (knowledge and skill) outcomes, with preliminary Level 3 evidence they may change learning behaviour. There was Level 2 evidence that a two-day dementia interprofessional “bootcamp” incorporating a range of learning methods (interactive, simulation, learning from lived experience) improved knowledge, confidence, and empathy for up to three months. While most studies evaluated interventions that aligned with DCSETF Tier 2, some aimed to increase awareness (Tier 1). Dementia Friends awareness courses (in-person or online) improved attitudes and knowledge immediately (Kirkpatrick Level 2). By contrast, asynchronous, self-directed learning did not improve learning outcomes, relative to standard training.

Most of the evidence we identified was at Kirkpatrick Level 2 (improved learning outcomes); only two studies provided evidence on change in behaviour or practice (Kirkpatrick Level 3) and none of effects on patient outcomes (Kirkpatrick Level 4). Though almost all training programmes sought to learn from lived experience, none consulted patients regarding the impact of the training on students’ skills and practice. Successful learning may not necessarily lead to successful practice change; other factors, including an enabling workplace culture, strong clinical leadership and self-motivation are also required^53^. Future studies might usefully consider patient perspectives in determining the impact of educational programmes.

As with the earlier ‘*What Works*’ review^14^, this review did not find high quality evidence that asynchronous, self-directed learning enhances knowledge or improves attitudes, relative to standard training. Surr et al point to the importance of integration of classroom and patient-based learning^14^. Additionally, we found that greater integration in clinical training could be beneficial. This includes integrating skills-based learning within clinical placements and promoting interprofessional learning. Where undergraduates from different disciplines learned together^37,49^ or with care staff^27^ it was generally valued and successful.

The initial findings of the Darzi review note the urgent need for more consistent training; undergraduate training programmes are an opportunity to deliver this^54^. Effective interventions increased confidence and enjoyment of dementia care encounters, and increased interest in careers in dementia specialties^20,27^. Notable absences in evidence for undergraduate training related to occupational therapy, physiotherapy, clinical pharmacy, social work and other professional groups. As noted by our stakeholder group, learning during placements relies on the skills and knowledge of staff, underlining the importance of quality dementia education for all healthcare workers. Attracting and supporting a workforce who are motivated to pursue dementia care and have appropriate training to provide good quality care to people with dementia in non-dementia-specialist settings (such as primary care, general hospitals, and social care) is key to delivering good quality future care. Mandating evidence-based skills programmes across specialties to ensure that students learn the skills and competencies required to be part of an effective future workforce could drive important improvements in future care quality.

## Data Availability

No data was produced.

## Acknowledgements

This research is funded through the NIHR Policy Research Unit in Dementia and Neurodegeneration – Queen Mary University of London (reference NIHR206110). The views expressed are those of the author(s) and not necessarily those of the NIHR or the Department of Health and Social Care.

