## Supplementary material for "Dementia Education and Training for the Multidisciplinary Student Healthcare Workforce: A Systematic Review"

**Table S1: Search strategy for PubMed (dates 2015- filter applied)**

| Terms |  |
| --- | --- |
| Education | (educat*[Title/Abstract] OR training[Title/Abstract] OR staff development[Title/Abstract] OR professional development[Title/Abstract] OR CPD[Title/Abstract] OR skills training[Title/Abstract] OR curricul*[Title/Abstract] OR learn*[Title/Abstract] OR teach*[Title/Abstract] OR workshop[Title/Abstract] OR module[Title/Abstract]) |
| Staff | ((professional[Title/Abstract] OR staff[Title/Abstract] OR worker[Title/Abstract] OR workforce[Title/Abstract] OR paid carer[Title/Abstract] OR aide[Title/Abstract] OR care worker[Title/Abstract] OR physician[Title/Abstract] OR doctor[Title/Abstract] OR student[Title/Abstract] OR nurse[Title/Abstract] OR therapist[Title/Abstract] OR social worker[Title/Abstract]) |
| Dementia | (dementia[Title/Abstract] OR "Frontotemporal dementia"[Title/Abstract] OR korsakof*[Title/Abstract] OR binswanger[Title/Abstract] OR "Progressive Supranuclear palsy"[Title/Abstract] OR alzheimer*[Title/Abstract] OR dement*[Title/Abstract] OR "Korsakoff Syndrome"[Title/Abstract] OR "Wernicke's Encephalopathy"[Title/Abstract] OR "Huntington's Disease"[Title/Abstract] OR "Multi Infarct"[Title/Abstract] OR "lewy bodies"[Title/Abstract] OR "Lewy Body Disease"[Title/Abstract] OR "kluver-bucy syndrome"[Title/Abstract] OR "Vascular dementia"[Title/Abstract] OR "Creutzfeldt-Jakob Syndrome"[Title/Abstract] OR "Alzheimer's Disease"[Title/Abstract])) |

**Table S2: Quality rating of studies for healthcare workers in training, using the MMAT**

|  | ***Qualitative*** | | | | | ***Quantitative non-randomised*** | | | | | ***Quantitative descriptive*** | | | | | ***Mixed Methods*** | | | | |
| --- | --- | --- | --- | --- | --- | --- | --- | --- | --- | --- | --- | --- | --- | --- | --- | --- | --- | --- | --- | --- |
|  | 1.1 | *1.2* | *1.3* | *1.4* | *1.5* | *3.1* | *3.2* | *3.3* | *3.4* | *3.5* | *4.1* | *4.2* | *4.3* | *4.4* | *4.5* | *5.1* | *5.2* | *5.3* | *5.4* | *5.5* |
| *Annear*^1^ | *x* | *x* | *x* | *x* | *x* | *x* | *x* | *x* | *-* | *x* |  |  |  |  |  | *x* | *x* | *x* | *x* | *x* |
| *Balzer*^2^ | *x* | *x* | *x* | *-* | *-* | *x* | *x* | *x* | *-* | *x* |  |  |  |  |  | *-* | *x* | *x* | *x* | *-* |
| *Banerjee*^3^ |  |  |  |  |  | *x* | *x* | *-* | *x* | *x* |  |  |  |  |  |  |  |  |  |  |
| *Bard* ^4^ |  |  |  |  |  | *x* | *-* | *x* | *-* | *x* |  |  |  |  |  |  |  |  |  |  |
| *Berning*^5^ |  |  |  |  |  | *x* | *x* | *x* | *-* | *x* |  |  |  |  |  |  |  |  |  |  |
| *Brown*^6^ | *x* | *x* | *x* | *x* | *x* | *x* | *x* | *-* | *-* | *x* |  |  |  |  |  | *-* | *x* | *x* | *x* | *-* |
| *Craig*^7^ |  |  |  |  |  | *x* | *x* | *x* | *-* | *-* |  |  |  |  |  |  |  |  |  |  |
| *Daley*^8^ | *x* | *x* | *x* | *x* | *x* |  |  |  |  |  |  |  |  |  |  |  |  |  |  |  |
| *Daley*^9^ |  |  |  |  |  | *x* | *x* | *-* | *x* | *-* |  |  |  |  |  |  |  |  |  |  |
| *Davison*^10^ | *x* | *x* | *x* | *x* | *x* |  |  |  |  |  | *x* | *-* | *x* | *-* | *-* | *-* | *x* | *x* | *x* | *-* |
| *DeCaro*^11^ |  |  |  |  |  | *x* | *x* | *-* | *-* | *x* |  |  |  |  |  |  |  |  |  |  |
| *Dellasega*^12^ | *x* | *x* | *x* | *x* | *x* |  |  |  |  |  |  |  |  |  |  |  |  |  |  |  |
| *Dingwall et al. (2017)* | *-* | *x* | *x* | *-* | *x* | *x* | *-* | *x* | *-* | *x* |  |  |  |  |  | *x* | *x* | *-* | *x* | *-* |
| *Dressel et al. (2023)* | *x* | *x* | *x* | *-* | *-* | *x* | *x* | *-* | *-* | *x* |  |  |  |  |  | *-* | *x* | *-* | *x* | *-* |
| *Frausing*^13^ | *x* | *x* | *x* | *x* | *x* |  |  |  |  |  |  |  |  |  |  |  |  |  |  |  |
| *Griffiths*^14^ |  |  |  |  |  | *x* | *x* | *-* | *-* | *x* |  |  |  |  |  |  |  |  |  |  |
| ***Grosvenor***^15^ | ***x*** | ***x*** | ***x*** | ***x*** | ***x*** |  |  |  |  |  |  |  |  |  |  |  |  |  |  |  |
| *Harrington*^11^ | *x* | *x* | *x* | *x* | *x* | *-* | *-* | *x* | *-* | *x* |  |  |  |  |  | *x* | *x* | *-* | *-* | *-* |
| *Hartung*^16^ |  |  |  |  |  | *x* | *x* | *-* | *-* | *x* |  |  |  |  |  |  |  |  |  |  |
| *Haugland*^17^ | *x* | *x* | *x* | *x* | *x* |  |  |  |  |  |  |  |  |  |  |  |  |  |  |  |
| *Jones*^18^ |  |  |  |  |  | *x* | *x* | *-* | *-* | *x* |  |  |  |  |  |  |  |  |  |  |
| *Long*^19^ |  |  |  |  |  | *-* | *x* | *x* | *-* | *-* |  |  |  |  |  |  |  |  |  |  |
| *Love*^20^ |  |  |  |  |  | *x* | *x* | *x* | *x* | *x* |  |  |  |  |  |  |  |  |  |  |
| *Mastel-Smith*^21^ | *x* | *x* | *x* | *x* | *x* | *-* | *x* | *x* | *-* | *x* |  |  |  |  |  | *x* | *x* | *x* | *x* | *-* |
| *Mosley*^22^ |  |  |  |  |  | *x* | *x* | *-* | *-* | *x* |  |  |  |  |  |  |  |  |  |  |
| *Naughton et al*^23^ | *x* | *x* | *x* | *x* | *x* |  |  |  |  |  |  |  |  |  |  |  |  |  |  |  |
| *Naughton*^24^ | *x* | *x* | *x* | *x* | *x* | *x* | *x* | *-* | *-* | *x* |  |  |  |  |  | *-* | *x* | *x* | *x* | *-* |
| *Niedens*^25^ | *x* | *x* | *x* | *x* | *x* | *x* | *x* | *-* | *-* | *x* |  |  |  |  |  | *x* | *x* | *x* | *x* | *-* |
| *Peng*^26^ | *x* | *x* | *x* | *-* | *x* | *x* | *x* | *x* | *-* | *-* |  |  |  |  |  | *-* | *x* | *x* | *x* | *-* |
| *Salajegheh* ^27^ | *x* | *x* | *x* | *x* | *x* |  |  |  |  |  |  |  |  |  |  |  |  |  |  |  |
| *Schwarz*^28^ |  |  |  |  |  | *-* | *x* | *x* | *-* | *x* |  |  |  |  |  |  |  |  |  |  |
| *Smyth*^29^ | *x* | *x* | *x* | *x* | *x* | *-* | *-* | *-* | *x* |  |  |  |  |  |  | *-* | *x* | *-* | *-* | *-* |
| *Um*^30^ | *x* | *x* | *x* | *x* | *x* |  |  |  |  |  |  |  |  |  |  |  |  |  |  |  |
| *Winter*^31^ | *-* | *x* | *-* | *x* | *-* |  |  |  |  |  |  |  |  |  |  |  |  |  |  |  |
| *Wonnacott*^32^ | *x* | *x* | *x* | *x* | *x* |  |  |  |  |  |  |  |  |  |  |  |  |  |  |  |

***‘X’*** = criteria met; **‘-‘** = criteria not met

**MMAT, 2018 methodological quality criteria:**

**1. Qualitative;** 1.1. Is the qualitative approach appropriate to answer the research question? 1.2. Are the qualitative data collection methods adequate to address the research question? 1.3. Are the findings adequately derived from the data? 1.4. Is the interpretation of results sufficiently substantiated by data? 1.5. Is there coherence between qualitative data sources, collection, analysis and interpretation?

**3. Quantitative non-randomised;** 3.1. Are the participants representative of the target population? 3.2. Are measurements appropriate regarding both the outcome and intervention (or exposure)? 3.3. Are there complete outcome data? 3.4. Are the confounders accounted for in the design and analysis? 3.5. During the study period, is the intervention administered (or exposure occurred) as intended?

**4. Quantitative descriptive;** 4.1. Is the sampling strategy relevant to address the research question? 4.2. Is the sample representative of the target population? 4.3. Are the measurements appropriate? 4.4. Is the risk of nonresponse bias low? 4.5. Is the statistical analysis appropriate to answer the research question?

**5. Mixed Methods;** 5.1. Is there an adequate rationale for using a mixed methods design to address the research question? 5.2. Are the different components of the study effectively integrated to answer the research question? 5.3. Are the outputs of the integration of qualitative and quantitative components adequately interpreted? 5.4. Are divergences and inconsistencies between quantitative and qualitative results adequately addressed? 5.5. Do the different components of the study adhere to the quality criteria of each tradition of the methods involved? Note: 5.5 was rated ‘Yes’ if individual Qualitative and Quantitative components were rated 4+.

**Table S3: Characteristics of studies rated <4 on the MMAT appraisal tool**

| **EXPERIENTIAL INTERVENTIONS INVOLVING VISITS OR SHARED ACTIVITIES WITH PEOPLE WITH DEMENTIA (N=3)** | | | | | | | | | | | | | |
| --- | --- | --- | --- | --- | --- | --- | --- | --- | --- | --- | --- | --- | --- |
| **Study**  **Country** | **Settings and Participants** | **Intervention** | **n** | **Control** | **n** | **n (%) with primary outcome)** | **Outcome (from baseline)** | **Outcomes: Kirkpatrick levels** | | | | **Study type** | **Validity score** |
|  |  |  |  |  |  |  |  | **1** | **2** | **3** | **4** |  |  |
| Daley^33^  UK | Healthcare students from 5 universities | TFD Programme: students visit a PLWD and their family carer (See Table 1) | 2700 | UT | 863 | 3312 (93%) | 24 months |  | ☺ |  |  | Quant | 3/5 |
| Mosley^22^  USA | Pharmacy students from 1 university | 3 outreach sessions where the students were paired with a PLWD to encourage fidget blanket use through patient-specific directions and demonstration. | 26 | _ | _ | 12/26 (46%) | PI^e^ |  | ☺ |  |  | Quant | 3/5 |
| Balzer^2^ Germany | Nursing and medical students | Lectures, problem-based learning tutorials and visits to care facilities. Contact hours with lecturers, tutors or healthcare representatives around 30 hours. | 18 | _ | _ | 18/18 (100%) | PI | ☺ | ☺ |  |  | MM | 3/5 |
| **SKILLS TRAINING DURING CLINICAL PLACEMENT (N=3)** | | | | | | | | | | | | | |
| Hartung^16^  Canada | 1^st^ year nursing students completing clinical placements at geriatric healthcare settings | 2.75-hour workshop that applied a person-centred communication framework when caring for PLWD experiencing responsive behaviours during the students' 13-week clinical placements. | 43 | - | - | 33/43 (76.7%) | PI and 10-week follow-up |  |  |  | ☺ | Quant | 3/5 |
| Naughton^23^ UK | Pre-registration student nurses in 5 older adult units across two hospitals | 2.5-hour training sessions using VERA framework communication tool, plus reflective discussions, in placement | 51 | UT | 66 | 52/117 (44%) | PI | ☺ | ☺ |  |  | MM | 3/5 |
| Smyth^29^  Ireland | 2^nd^ year undergraduate nursing students in a residential facility | 2.5-hour, in-person VERA communication skills training by two trained researchers, strategies used during placement. | 10 | UT | 6 | 6/6 (100%) | PI |  | ☺ |  |  | MM | 1/5 |
| **SELF-DIRECTED ONLINE LEARNING (N=3)** | | | | | | | | | | | | | |
| Brown^6^ USA | Pre-clinical nursing students from 3 nursing schools in Connecticut and Hawaii | Modular, flipped classroom curriculum showing character animation techniques and videos teaching cognition and dementia, respectively (2 sessions). | 223 | - | - | 152/223 (68%) | PI | ☺ | ☺ |  |  | MM | 3/5 |
| Craig^7^  Northern Ireland | 1^st^ year undergraduate nursing students | Digital game with multiple-choice questions about dementia. Game about 90 seconds, can be played multiple times. | 452 | - | - | 334 (73.9%) | PI |  | ☺ |  |  | Quant | 3/5 |
| Long^19^  USA | Nursing students from Lamar University | Four online, interactive educational modules, focused on caring for PLWD, clinical reasoning abilities and student confidence through video vignettes. | 65 | - | - | 65/65 (100%) | PI |  | ☺ |  |  | Quant | 2/5 |
| **DEMENTIA AWARENESS (DEMENTIA FRIENDS) TRAINING (N=2)** | | | | | | | | | | | | | |
| Griffiths^14^  Malaysia | Pharmacy and medical undergraduate students from 1 University | One hour dementia education; dispels myths around dementia; demonstration of content, interactive activities in 5 teams, videos, discussion, case study | 112 | - | - | 97/112 (86.6%) | PI |  | ☺ |  |  | Quant | 3/5 |
| Davison^10^ UK | First-year healthcare students (medicine, nursing, speech and language therapy, OP, physiotherapy, pharmacy, paramedic science) from University of East Anglia | Inter-professional team created a learning package for students to gain a Tier 1 DA^h^ qualification, using the platform of an existing inter professional learning module. It comprised of 1-hour DF^i^ session (face-to-face or online), a reflective workbook and 10 multiple choice questions whereby 8/10 was required for the DA qualification. | 60 | - | - | 57/60 (95%) | PI | ☺ | ☺ |  |  | MM | 3/5 |
| **EXPERIENCING AND REFLECTING ON SIMULATION/FICTIONAL SCENARIOS (N=6)** | | | | | | | | | | | | | |
| Harrington^11^  USA | Nurse practitioner students from one university and one nursing college | VDT^j^: Trained facilitators guide students through an eight-minute experience of a PLWD perspective, as they try to perform five daily activities, followed by a 45-minute focus group. | 44 | - | - | 20/44 (45.5%) | PI |  | ☺ |  |  | MM | 2/5 |
| Schwarz^28^  USA | 1^st^ year DPT^k^ students at a local memory care facility | 2-hour experimental learning activity involving a brief, simulated dementia experience, educational session, observational facility tour and a debriefing session. Facilitated by dementia-training educators | 82 | - | - | 80/82 (95.4%) | PI and nine-month follow-up |  | ☺ |  |  | Quant | 3/5 |
| Dingwall^34^ UK | 3^rd^ year nursing and social work students from one university | ‘Sliding Doors’, drama-based, educational one-day workshop; dramatized scenarios to stimulate discussions. | 63 | - | - | Unclear | PI | ☺ | ☺ |  |  | MM | 3/5 |
| Bard^4^  USA | 2^nd^ year medical students | 30-minute VR simulation session, through the perspective of a PLWD. | 149 | - | - | 149/149 (100%) | PI |  | ☺ |  |  | Quant | 3/5 |
| Peng^26^  China | 2^nd^ year undergraduate nursing students | Movie overview of ‘Still Alice’, about a PLWD; 8-minute, 5-task modified VDT. | 45 | - | - | N/A | PI | ☺ | ☺ | ☺ |  | MM | 3/5 |
| Winter^31^  UK | All 3^rd^ year medical students from one university | 12 simulation days over 9 months, based on communicating with a person with dementia in several scenarios facilitated by doctors, nurses and occupational therapists specialised in dementia care. Students managed simulated scenarios in pairs, followed by a 30-minute debrief. | 145 | - | - | N/A | PI | ☺ | ☺ |  |  | Qual^m^ | 2/5 |
| **CLASSROOM-BASED LEARNING (N=3)** | | | | | | | | | | | | | |
| Jones^18^  UK | Student paramedics from 1 university | A 6-hour education program with didactic teaching, collaborative and reflective learning. Face to face by specialist dementia educator; involved quizzes, case studies, videos; prerequisite learning | 43 | - | - | 32/43 (74.4%) | PI | ☺ | ☺ |  |  | Quant | 3/5 |
| DeCaro^35^ USA | Medical students, undergraduate and graduate students interested in medicine | 6 educational and 3 outreach events; monthly dementia focused didactic meetings and outreach focusing on Black participant recruitment | 37 | _ | _ | 20/37 (54%) | PI |  | ☺ |  |  | Quant | 3/5 |
| Dressel ^36^et al. (2023)  Germany | Nutrition therapy and counselling, speech language pathology and physiotherapy students from 1 University | Interprofessional and competency-based education; three-day workshop focusing on patient-centred care and inter professional collaboration; case-based leaning in simulated interprofessional case-conferences and peer teaching. | 42 | - | - | 42/53 (79.2%) | PI |  | ☺ |  |  | MM | 2/5 |

**Legend**: DA: Dementia Awareness; DF: Dementia Friends; DPT: Doctor of Physical Therapy; MM: mixed methods; PI: post-intervention; PLWD: person living with dementia; Qual: qualitative; Quant: quantitative; TFD: Time for Dementia; UT: Usual Training; VERA: Validation, Emotion, Reassurance, Activity; VDT: Virtual Dementia Tour; VR: Virtual Reality;
